# Investigating the DNA methylation profile of e-cigarette use

**DOI:** 10.1101/2021.01.28.21250699

**Authors:** Rebecca C Richmond, Carlos Sillero Rejon, Jasmine N Khouja, Claire Prince, Alexander Board, Gemma Sharp, Matthew Suderman, Caroline L Relton, Marcus Munafò, Suzanne H Gage

## Abstract

**Rationale and objectives:** Little evidence exists on the health effects of e-cigarette use. DNA methylation may serve as a biomarker for exposure and could be predictive of future health risk. We aimed to investigate the DNA methylation profile of e-cigarette use.

**Methods:** Among 117 smokers, 117 non-smokers and 116 non-smoking vapers, we evaluated associations between e-cigarette use and epigenome-wide methylation from saliva. We tested associations between e-cigarette use and methylation scores known to predict smoking and smoking-related disease. We assessed the ability of a methylation score for predicting e-cigarette use and for discriminating lung cancer.

**Measurements and Main Results:** 7 CpGs were identified in relation to e-cigarette use at p<1×10^−5^ and none at p<5.91×10^−8^. 13 CpGs were associated with smoking at p<1×10^−5^ and one at p<5.91×10^−8^. CpGs associated with e-cigarette use were largely distinct from those associated with smoking. There was strong enrichment of known smoking-related CpGs in the smokers but not the vapers. A methylation score for e-cigarette use showed poor prediction internally (AUC 0.55, 0.41-0.69) and externally (AUC 0.57, 0.36-0.74) compared with a smoking score (AUCs 0.80) and was less able to discriminate lung squamous cell carcinoma from adjacent normal tissue (AUC 0.64, 0.52-0.76 versus AUC 0.73, 0.61-0.85).

**Conclusions:** The DNA methylation profile for e-cigarette use is largely distinct from that of cigarette smoking, did not replicate in independent samples, and was unable to discriminate lung cancer from normal tissue. The extent to which methylation related to long-term e-cigarette use translates into chronic effects requires further investigation.

**Key Messages:** *What is the key question?:* Is there a DNA methylation signature of e-cigarette use and is it distinct from that of smoking?

*What is the bottom line?:* Smoke exposure is known to lead to widespread changes in DNA methylation which have been identified in different populations and samples, persist for many years after smoking cessation, and may act as a biomarker for smoking-related disease risk and mortality. Whether a similar methylation profile exists in relation to e-cigarette use has not been widely investigated.

*Why read on?:* We obtained saliva samples from 116 e-cigarette users and compared their DNA methylation profile with 117 smokers and 117 non-smokers. The e-cigarette users in this study had a minimal smoking history, and so we were able to distinguish the effects of e-cigarette use from those of smoke exposure. Overall, we found that the methylation profile associated with e-cigarette use is less pronounced and distinct from that associated with cigarette smoking.

## Introduction

Electronic cigarettes (e-cigarettes) have the potential to reduce smoking-related harm. Although little evidence currently exists on long-term effects, their lack of tar and very low levels of other dangerous substances^1^ suggest they are considerably less harmful than cigarettes.^2^ They have been shown to be an efficacious^3^ and cost-effective^4^ smoking cessation aid. While it will take years to fully estimate the impact of e-cigarette use on diseases including cancer, we can investigate whether it is associated with any biomarkers that may predict future health risk.^5^

Recent studies have found a reduction in harmful biomarkers among e-cigarette users compared with smokers, with some biomarkers showing levels similar to non-smokers.^5-7^ However, only a few biomarkers have been investigated and all with relatively short half-lives,^8 9^ meaning their utility for predicting long-term effects of e-cigarette use may be limited. DNA methylation is a type of epigenetic modification involving the addition of methyl groups to the DNA which influences how the underlying DNA sequence is interpreted and expressed. Pronounced differences in DNA methylation have been found between cigarette smokers and non-smokers,^10^ have been replicated in different populations^11 12^ and tissues,^13^ have been shown to persist for several years post-cessation,^10^ and have been shown to distinguish tumour from normal samples ^14^. DNA methylation profiles can act as a biomarker of tobacco smoke exposure, can be used to categorise participants in relation to smoking status, and are predictive of disease and mortality.^15 16^

It has been proposed that the products of tobacco combustion might be responsible for the change in methylation rather than tobacco itself, since no distinctive methylation patterns associated with smokeless tobacco (“snuff”) have been identified.^25^ However, this previous study only investigated peripheral blood and not tissue-specific methylation changes at the site of exposure (i.e. oral samples). Support for the investigation of methylation in saliva has recently been provided given the overlap in methylation signals related to smoke exposure in both blood and saliva^17^, with one study demonstrating a stronger signal in saliva compared with matched blood samples^27^.

Assessing the DNA methylation profile of e-cigarette users could inform our understanding of the potential biological impact associated with their use and the relative health risks compared to cigarettes.^18^ We propose that saliva is the most appropriate sample type to conduct an investigation into the potential impact of e-cigarette use on DNA methylation, as it represents the location of direct exposure-tissue contact and is a non-invasive sample which can be easily obtained.

The SEE-Cigs study (<u>S</u>tudying the <u>E</u>pigenetics of <u>E-cig</u>arette Use) was set up to recruit a sample of vapers, smokers and non-smokers, from whom we obtained detailed questionnaire data on smoking and vaping habits, demographic characteristics, as well as saliva samples from which DNA methylation was assessed. In the present study, we explored whether e-cigarette use is associated with DNA methylation and evaluated the degree of similarity between DNA methylation profiles in e-cigarette users (vapers) and cigarette smokers (compared with non-smokers) in saliva samples. We investigated associations between e-cigarette use and previously-developed DNA methylation scores used to predict smoking-related disease and mortality,^12 16^ and biological ageing.^19 20^ We also generated a novel DNA methylation score for predicting e-cigarette use and investigated this in lung tumour and adjacent normal tissue, in order to make inferences about the potential importance of e-cigarette-related DNA methylation in lung cancer development.

## Methods

The analysis plan was pre-registered^21^ and is summarized in **Supplementary Figure 1**.

**Figure 1:**
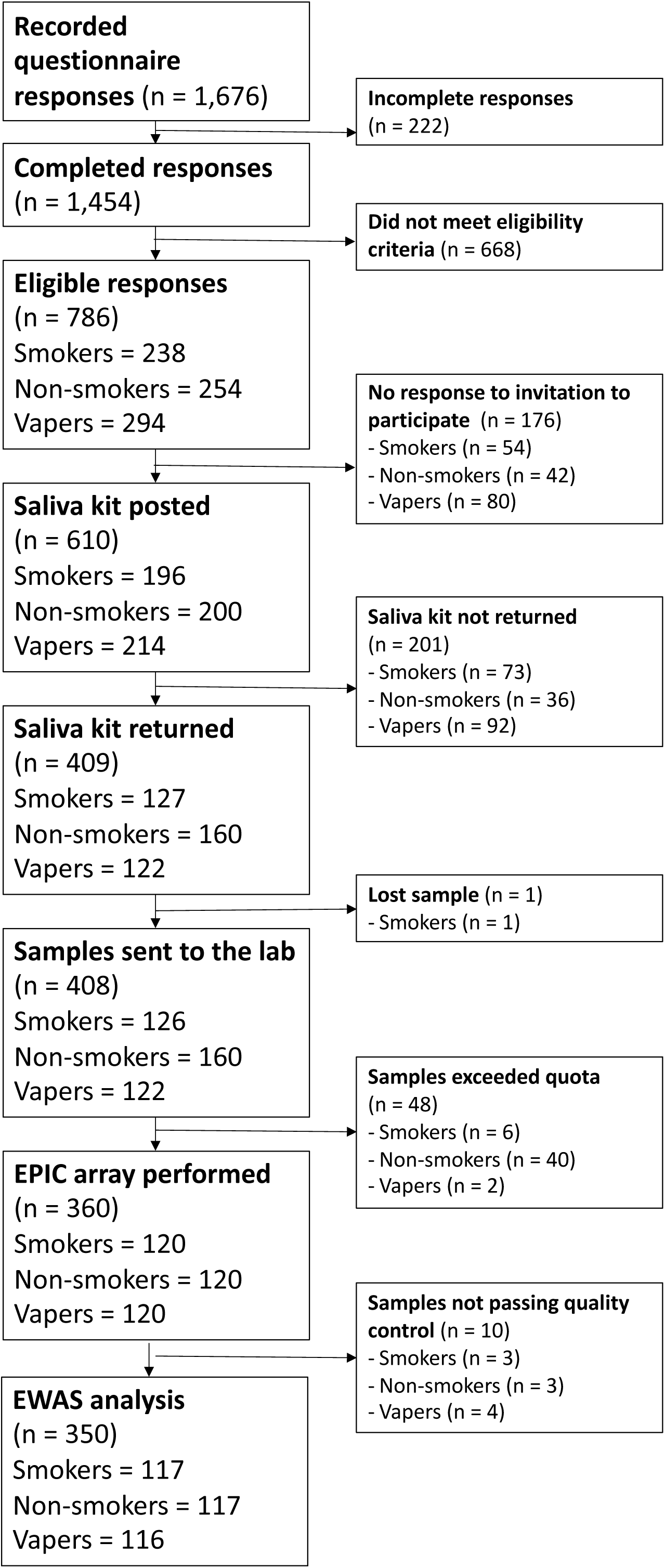
Participant flow chart for SEE-Cigs

### Study Design

The SEE-Cigs study (<u>S</u>tudying the <u>E</u>pigenetics of <u>E-cig</u>arette Use) recruited vapers, smokers and non-smokers from the United Kingdom general population. It was important that vapers did not have a long previous smoking history, given the persistence of DNA methylation marks associated with smoke exposure many years after cessation^10 22^ Vapers were therefore defined as having used e-cigarettes at least weekly for the past 6 months and having smoked <100 times in their lifetime; smokers as having smoked at least weekly for the past 6 months and having used an e-cigarette <100 times in their lifetime; never smokers as having smoked and/or used an e-cigarette <100 times in their lifetime. We aimed to recruit 120 participants per group (vapers, smokers, non-smokers) in order to provide >80% power to detect a 4.5% mean difference in DNA methylation at p<0.05 and >80% power to detect an 11% mean difference at p<1×10^−6^. Further eligibility criteria and recruitment details are described in the **Online Supplement**.

### Questionnaire

We asked participants questions about their smoking and vaping behavior, as well as socio-demographic and behavioural factors including age, gender, height and weight, ethnicity, education, occupation, household smoking, and recreational drug use. Participants were allocated to three participant groups (smokers, vapers and never-smokers), after which additional group-specific questions were asked (see **Online Supplement**).

### DNA methylation

Oragene™ kits (www.dnagenotek.com/US/products/OGR500.html) were used for collecting saliva samples. Genome-wide methylation status of over 850,000 CpGs was measured using the Illumina HumanMethylationEPIC array according to standard protocol. Details of sample acquisition, processing, data quality control and normalization are provided in the **Online Supplement**.

## Statistical analysis

### Epigenome-wide association study (EWAS)

Multivariable linear regression was used to assess the differences in DNA methylation at each measured CpG site between (1) vapers vs. non-smokers, (2) smokers vs. non-smokers, and (3) smokers vs. vapers, with adjustment for age, biological sex, body mass index (BMI), educational attainment, household smoking, recreational drug use and 20 surrogate variables, using *meffil*.^23^ We investigated CpGs which reached a Bonferroni-significance threshold of p<5.91×10^−8^ (0.05/846,244 CpGs tested), as well as a less stringent threshold of p<1×10^−5^. From these EWAS results, we identified differentially methylated regions (DMRs) using the *dmrff* R package.^24^ DMRs were defined as regions containing at least two CpGs within 500 bp, each with EWAS meta-analysis P-values <0.05 and methylation changes in a consistent direction, and where the regional p-value surpassed Bonferroni correction. For the EWAS of vapers vs. non-smokers, three additional models were run: i) with adjustment for estimated cell type composition (see **Online Supplement**), ii) with adjustment for previous smoking history (number of cigarettes) and iii) restricted to individuals of white ethnicity.

### Enrichment and annotation

From the EWAS results of (1) vapers vs. non-smokers, and (2) smokers vs. non-smokers, we investigated evidence for enrichment of associations among 2,623 and 1,501 smoking-related DNA methylation sites identified in previous large-scale studies of blood (Joehanes et al, 2016)^10^ and buccal samples (Teschendorff et al, 2015),^14^ using a Wilcoxon rank sum test.

While we excluded potential participants who reported drug dependence, given the widespread use of e-cigarettes for inhaling cannabinoids^25^, we assessed whether any of the 15 CpG sites identified in an EWAS of cannabis^26^ were associated with e-cigarette use after Bonferroni correction. We also investigated whether there was any evidence for replication of 14 CpGs related to e-cigarette use in a previous EWAS,^27^ and assessed the extent to which the CpGs identified in our EWAS had been previously reported in relation to other traits in two publicly available repositories.^28 29^ We explored the potential functions of the top 50 CpGs identified in each EWAS via GO and KEGG enrichment analysis using the *missMethyl* R package.^30^

### DNA methylation scores for smoking and epigenetic ageing

DNA methylation scores can be derived by summing methylation values at relevant CpG sites previously identified in relation to a relevant exposure, weighted by the effect sizes observed in independent epigenetic studies. Five DNA methylation scores of smoke exposure (Joehanes, McCartney, Teschendorff, Lu and *AHRR*) ^10 14 16 31 32^ and four DNA methylation scores of ageing (intrinsic epigenetic age acceleration (IEAA), extrinsic epigenetic age acceleration (EEAA), PhenoAge and GrimAge)^31 33-35^ were quantified. Details of the scores are provided in the **Online Supplement**.

Multivariable linear regression was used to assess differences in DNA methylation scores between the three groups with adjustment for age, sex, BMI, educational attainment, household smoking and recreational drug use. Further analyses were restricted to individuals of white ethnicity only, with adjustment for DNA methylation-derived smoking pack-years in the GrimAge model^31^, and with adjustment for self-reported smoking history when evaluating DNA methylation scores in relation to e-cigarette use.

### DNA methylation score for e-cigarette use

We generated DNA methylation scores for e-cigarette use and smoking within SEE-Cigs and then assessed the discriminative performance of these scores for predicting e-cigarette use and smoking within SEE-Cigs and in an independent dataset, the Avon Longitudinal Study of Parents and Children (ALSPAC)^36-38^. We also investigated whether the DNA methylation score for e-cigarette use was able to discriminate tumour from normal tissue in lung to the same extent as the DNA methylation score for smoking, using data from publicly available DNA methylation data in the Cancer Genome Atlas (TCGA)^2^. Details of the scores generated and the data used are provided in the **Online Supplement**.

## Results

### Descriptive characteristics

**Figure 1** shows the participant flow for the SEE-Cigs study. The final sample consisted of 117 smokers, 117 non-smokers and 116 vapers with DNA methylation data (see **Online Supplement**). Descriptive characteristics are displayed in **Table 1**. Compared with non-smokers, vapers were more likely to have higher BMI, be male, have lower educational attainment and be more exposed to household smoke, while smokers were more likely to be male, have lower educational attainment, be more exposed to household smoke and to use drugs recreationally.

**Table 1:** Descriptive characteristics of participant groups in this study

|  | **Non-smokers**<br>**(N = 117)** | **Smokers**<br>**(N = 117)** | **Vapers**<br>**(N = 116)** | **Overall P for**<br>**difference** |
| --- | --- | --- | --- | --- |
| **Socio-demographic**<br>**factors** | **mean (SD)** | **mean (SD)** | **mean (SD)** |  |
| Age (years) | 20.6 (3.5) | 22.8 (4.8) | 20.9 (4.3) | 0.044 |
| BMI (kg/m<sup>2</sup>) | 22.6 (3.5) | 23.1 (4.2) | 26.1 (7.0) | <0.001 |
|  | **median (IQR)** | **median**<br>**(IQR)** | **median (IQR)** |  |
| Smoking (pack-years) | 0 (0, 0.0001) | 1.20 (0.38,<br>3.15) | 0.0013 (0.0006,<br>0.0027) | <0.001 |
|  | **% (n)** | **% (n)** | **% (n)** |  |
| Sex (% male) | 50.0 (60) | 50 (60) | 80 (73) | <0.001 |
| Ethnicity (% white) | 80 (96) | 89.1 (106) | 88.7 (102) | 0.075 |
| Education (% higher<br>education) | 86.7 (104) | 79.0 (94) | 60 (69) | <0.001 |
| Occupation (%<br>unemployed) | 4.2 (5) | 8.4 (10) | 21 (18.3) | 0.001 |
| Household smoking (%<br>yes) | 40 (48) | 67.5 (81) | 67.2 (78) | <0.001 |
| Recreational drug use (%<br>yes) | 5.8 (7) | 26.3 (31) | 9.3 (11) | <0.001 |
| **Estimated cell type**<br>**proportions (%)** | **mean (SD)** | **mean (SD)** | **mean (SD)** |  |
| Buccal | 48.3 (18.2) | 49.4 (18.5) | 45.4 (17.1) | 0.201 |
| Bcell | 1.7 (1.2) | 2.1 (1.8) | 2.2 (2.1) | 0.100 |
| CD4T | 0.2 (0.4) | 0.2 (0.5) | 0.2 (0.7) | 0.808 |
| CD8T | 0.001 (0.005) | 0.04 (0.44) | 0.03 (0.24) | 0.477 |
| Granulocytes | 47.3 (19.8) | 45.4 (20.8) | 49.1 (19.3) | 0.357 |
| Mono | 0.4 (0.7) | 0.5 (0.9) | 0.6 (1.0) | 0.307 |
| NK | 1.1 (1.4) | 1.5 (2.6) | 1.6 (2.3) | 0.183 |
| **Vaping characteristics** | **% (n)** | **% (n)** | **% (n)** |  |
| E-cigarette device | N/A | N/A |  | N/A |
| Cigalike (1st<br>generation) |  |  | 1.7 (2) |  |
| Tanks (2nd generation) |  |  | 9.4 (11) |  |
| Mods (3rd generation) |  |  | 67.3 (76) |  |
| Pods (4th generation) |  |  | 15.0 (17) |  |
| Other |  |  | 6.2 (7) |  |
| Frequency of use (%<br>daily) |  |  | 92.9 (105) |  |
| Device contains nicotine<br>(% yes) |  |  | 96.4 (108) |  |
| Increased nicotine concentration in the past (% yes) |  |  | 14.1 (16) |  |
|  | **mean (SD)** | **mean (SD)** | **mean (SD)** |  |
| Nicotine content (mg/ml) | N/A | N/A | 5.6 (4.7) | N/A |
| Liquid per day (ml) |  |  | 7.6 (7.6) |  |

Smokers were slightly older on average than non-smokers and vapers. The majority of participants were of white ethnicity, with a slightly higher proportion of non-white individuals in the group of non-smokers. Smokers had smoked for a median of 1.20 (IQR 0.38, 3.15) pack-years, while both non-smokers and vapers had a minimal smoking history. There were no differences in cell type proportions of the saliva samples obtained from the participants. Most vapers used e-cigarettes containing nicotine and vaped daily.

### EWAS

DNA methylation at 7 CpGs was associated with e-cigarette use (vs non-smoking) at p<1×10^−5^ and none at p<5.91×10^−8^ (**Table 2**). The top three CpGs were located in protein-coding genes for a ribonuclease P/MRP subunit (*RPP14*), an insulin-like growth factor receptor (*IGF1R*) and a gamma-aminobutyric acid (GABA) A receptor (*GABRP*). After accounting for cell composition, smoking history and ethnicity, associations at the 7 CpGs remained but were weakened, with the exception of signals at cg12435725 (*RPP14*), which strengthened with adjustment for smoking history (p=9.02×10^−8^), and cg02066693, which was stronger among individuals of white ethnicity (p=6.27×10^−7^) (**Supplementary Table 1**). DNA methylation at 13 CpGs was associated with smoking (vs non-smoking) at p<1×10^−5^ and one at p<5.91×10^−8^: cg05575921 located in *AHRR* encoding the aryl hydrocarbon receptor repressor (**Supplementary Table 2**). DNA methylation at 14 CpGs was associated with e-cigarette use (vs smoking) at p<1×10^−5^, with the top association also at *AHRR* (cg05575921) (**Supplementary Table 3**). DNA methylation at cg05575921 was 8.2% (95% CI 5.7, 10.5) lower in smokers compared with non-smokers, and 7.1% (95% CI 4.6, 9.6) lower in smokers compared with vapers. In differentially methylated region (DMR) analysis, 31 DMRs were identified in vapers vs non-smokers, 39 in smokers vs. non-smokers and 81 in vapers vs smokers. The top DMR identified between vapers and non-smokers comprised 9 CpGs in *MUC4*, with higher methylation in vapers compared with non-smokers (7.3% (95% CI 5.7, 9.0; p=4.13×10^−18^) (**Supplementary Table 4**).

**Table 2:** Differentially methylated CpG sites associated with e-cigarette use vs non-smoking

| **CpG site** | **Chromosome** | **Position** | **Gene Symbol** | **Beta** | **SE** | **P-value\*** |
| --- | --- | --- | --- | --- | --- | --- |
| **cg12435725** | 3 | 58293450 | *RPP14* | -<br>0.060 | 0.012 | $6.43 \times 10^{-7}$ |
| **cg02066693** | 15 | 99366135 | *IGF1R* | -<br>0.019 | 0.004 | $5.47 \times 10^{-6}$ |
| **cg14872828** | 5 | 170210761 | *GABRP* | -<br>0.035 | 0.007 | $2.77 \times 10^{-6}$ |
| **cg12734956** | 12 | 112181430 | *ACAD10* | -<br>0.013 | 0.003 | $5.89 \times 10^{-6}$ |
| **cg02934082** | 3 | 122705793 | *SEMA5B* | -<br>0.036 | 0.008 | $5.92 \times 10^{-6}$ |
| **cg00388391** | 1 | 18459578 | *IGSF21* | 0.025 | 0.006 | $8.38 \times 10^{-6}$ |
| **cg10440286** | 22 | 37771664 | *ELFN2* | 0.044 | 0.010 | $9.31 \times 10^{-6}$ |

Model adjusted for age, sex, body mass index, educational attainment, household smoking, recreational drug use and 20 surrogate variables. n=111 vapers, n=117 non-smokers ; P-values $< 1 \times 10^{-5}$

**Table 3:**
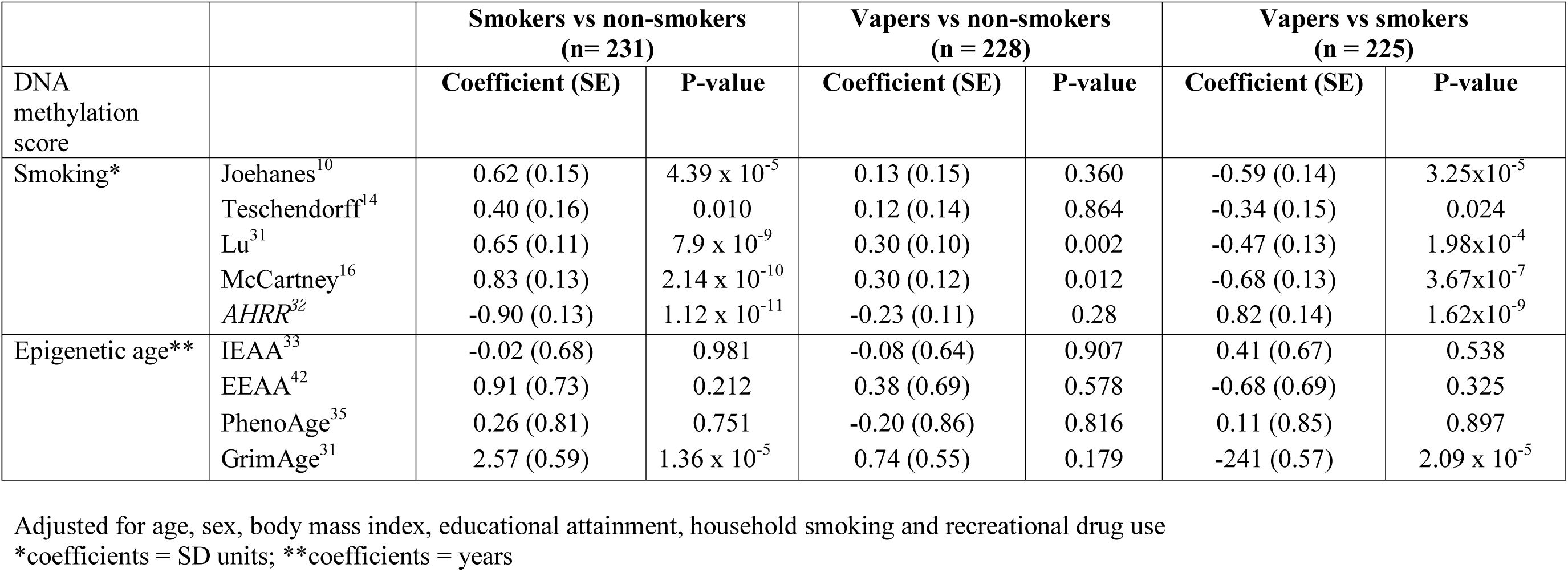
Differences in DNA methylation scores between participant groups

| DNA methylation score |  | Smokers vs non-smokers<br>(n= 231) |  | Vapers vs non-smokers<br>(n = 228) |  | Vapers vs smokers<br>(n = 225) |  |
| --- | --- | --- | --- | --- | --- | --- | --- |
|  |  | Coefficient (SE) | P-value | Coefficient (SE) | P-value | Coefficient (SE) | P-value |
| Smoking* | Joehanes<sup>10</sup> | 0.62 (0.15) | 4.39 x 10<sup>-5</sup> | 0.13 (0.15) | 0.360 | -0.59 (0.14) | 3.25x10<sup>-5</sup> |
|  | Teschendorff<sup>14</sup> | 0.40 (0.16) | 0.010 | 0.12 (0.14) | 0.864 | -0.34 (0.15) | 0.024 |
|  | Lu<sup>31</sup> | 0.65 (0.11) | 7.9 x 10<sup>-9</sup> | 0.30 (0.10) | 0.002 | -0.47 (0.13) | 1.98x10<sup>-4</sup> |
|  | McCartney<sup>16</sup> | 0.83 (0.13) | 2.14 x 10<sup>-10</sup> | 0.30 (0.12) | 0.012 | -0.68 (0.13) | 3.67x10<sup>-7</sup> |
|  | AHRR<sup>32</sup> | -0.90 (0.13) | 1.12 x 10<sup>-11</sup> | -0.23 (0.11) | 0.28 | 0.82 (0.14) | 1.62x10<sup>-9</sup> |
| Epigenetic age** | IEAA<sup>33</sup> | -0.02 (0.68) | 0.981 | -0.08 (0.64) | 0.907 | 0.41 (0.67) | 0.538 |
|  | EEAA<sup>42</sup> | 0.91 (0.73) | 0.212 | 0.38 (0.69) | 0.578 | -0.68 (0.69) | 0.325 |
|  | PhenoAge<sup>35</sup> | 0.26 (0.81) | 0.751 | -0.20 (0.86) | 0.816 | 0.11 (0.85) | 0.897 |
|  | GrimAge<sup>31</sup> | 2.57 (0.59) | 1.36 x 10<sup>-5</sup> | 0.74 (0.55) | 0.179 | -241 (0.57) | 2.09 x 10<sup>-5</sup> |

Adjusted for age, sex, body mass index, educational attainment, household smoking and recreational drug use
\*coefficients = SD units; \*\*coefficients = years

Apart from associations at *AHRR* in the models involving smoking, there was limited overlap in the top CpGs identified in the three EWAS (**Figure 2, Supplementary Figure 2**). 9 DMRs were found in common between at least two of the EWAS models (**Figure 2**). One DMR was hypermethylated (chr20:*BLCAP;NNAT*) and two hypomethylated (chr20:*SLC2A10* and chr3:*THRB*) in non-smokers compared with vapers and smokers. Two DMRs were hypermethylated (chr10 and chr3:*CACNA1D*) and two were hypomethylated (chr17:*BRCA1;NBR2* and chr6:*PRRT1;PPT2*) in smokers compared with non-smokers and vapers. Two DMRs were hypermethylated in vapers compared with smokers and non-smokers (chr10:*ANXA11;LINC00857* and chr17:*HSPB9;KAT2A*).

**Figure 2:**
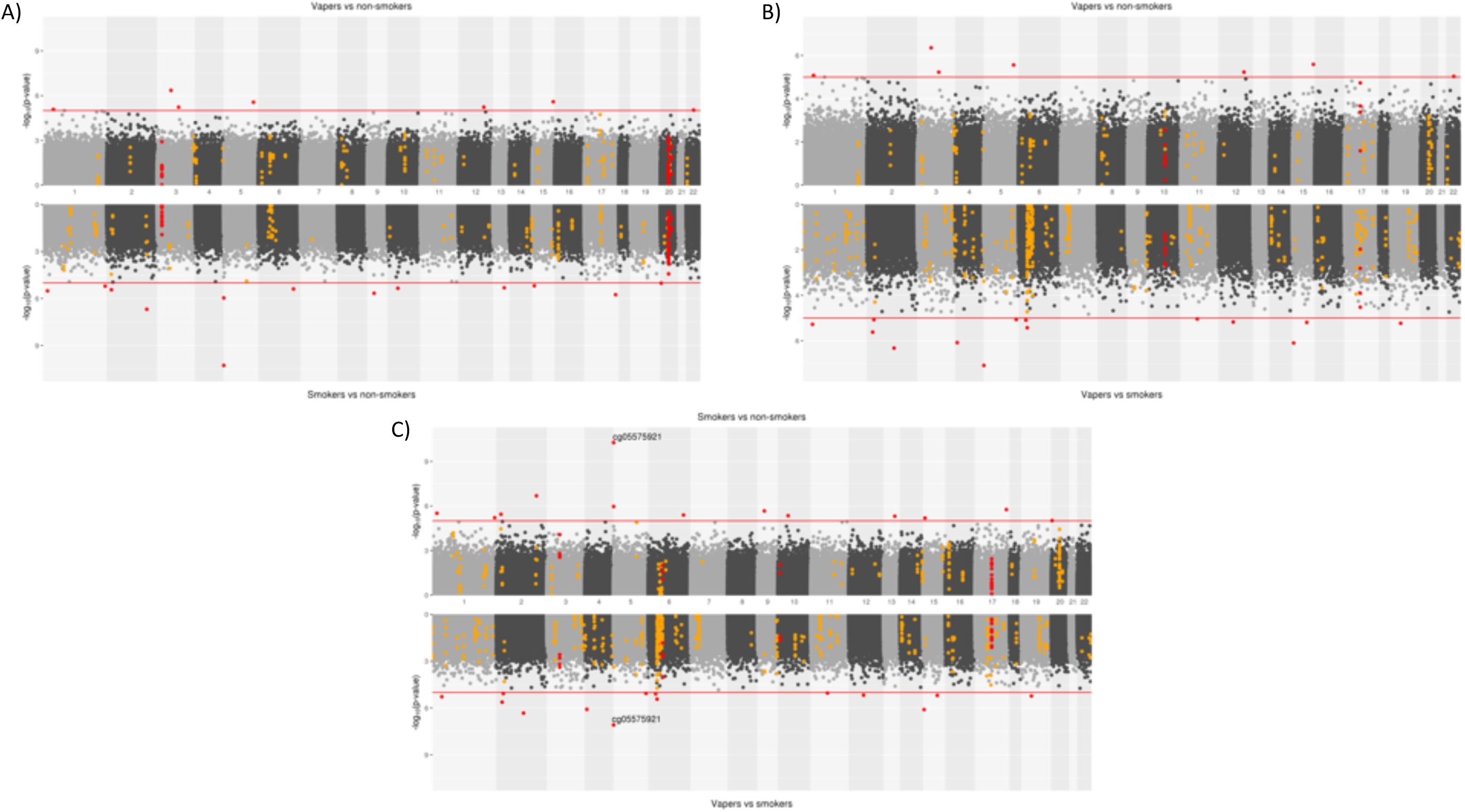
Comparison of epigenome-wide associations studies A) EWAS for e-cigarette use (vs. non-smoking) and smoking (vs. non-smoking) B) EWAS for e-cigarette use (vs. non-smoking) and e-cigarette use (vs. smoking) C) EWAS for smoking (vs. non-smoking) and e-cigarette use (vs. smoking)

### Enrichment and annotation

There was strong enrichment of known smoking-related CpGs in the EWAS of smokers vs. non-smokers (Wilcoxon test p=1.05×10^−14^ for Joehanes et al. CpGs^10^ and p=1.10×10^−9^ for Teschendorff et al. CpGs^14^) (**Supplementary Figure 2**), unlike in the EWAS of vapers vs. non-smokers (Joehanes p=0.67, Teschendorff p=0.28) (**Supplementary Figure 3**). One smoking-related CpG was found to be similarly differentially methylated in vapers in *GABRP* (cg14872828; p=2.77×10^−6^).

One CpG site previously associated with cannabis was found to be associated with e-cigarette use after Bonferroni correction (cg04180046; p=0.0029). This CpG has also been previously associated with smoke exposure^10^, and there was no difference in methylation at this site between vapers and smokers in the present study (p=0.865) (**Supplementary Table 5**).We also found little evidence of associations between 14 CpGs previously found in relation to e-cigarette use by Song et al.^27^ in any of the EWAS after Bonferroni correction (p>0.01) (**Supplementary Table 6**).

Three of the seven CpGs associated with e-cigarette use have been identified in previous EWAS for smoking, Down Syndrome, systemic corticosteroid, prostate cancer, gestational age and fetal vs. adult liver (**Supplementary Tables 7 and 8**). We found limited enrichment for KEGG pathways or GO terms (FDR p>0.05) (**Supplementary Tables 9-12**). In relation to e-cigarette use, response to ethanol/alcohol, positive regulation of insulin secretion and GABA transport were the top GO terms. Butanoate metabolism, synaptic vesicle cycle and GABAergic synapse were the top KEGG pathways.

### DNA methylation scores for smoking

All of the DNA methylation scores for smoking were correlated with reported pack-years smoked (|r|=0.21-0.52, p<0.0001), except the Teschendorff score^14^ (r=0.07, p=0.19) (**Supplementary Table 13**). All of the scores differed between the smokers vs. non-smokers (0.40-0.90 SD), and vapers vs. smokers (0.37-0.86 SD). While neither the Joehanes^10^ nor the Teschendorff score^14^ differed between vapers vs. non-smokers, higher levels of the other three scores (Lu,^31^ McCartney,^16^ *AHRR*) were observed in vapers vs. non-smokers (0.21-0.30 SD) (**Table 3**). These associations attenuated when reported smoking history was included in the model (**Supplementary Table 14**). Associations were similar in analyses restricted to individuals of white ethnicity (**Supplementary Table 15**).

### DNA methylation scores for epigenetic ageing

Epigenetic age estimates were strongly correlated with chronological age (r = 0.49-0.70, p<0.0001) (**Supplementary Table 16**). There was little difference in estimates of epigenetic age acceleration (EAA) between the three groups, except for GrimAge, where smokers had higher EAA relative to both vapers (2.5, 95% CI = 1.4-3.6 years) and non-smokers (2.6, 1.4-3.7 years) (**Table 3**). Associations persisted in analyses adjusting for a DNA methylation score for smoking pack-years^31^ (**Supplementary Table 17**) and when restricted to individuals of white ethnicity (**Supplementary Table 15**).

### Using DNA methylation as a biomarker of e-cigarette use and lung cancer

We generated DNA methylation scores for predicting e-cigarette use (**Supplementary Table 18**). These performed poorly at discriminating e-cigarette users from non-smokers in both the internal (AUC 0.55, 0.41-0.69) and external validation sets (AUC 0.57, 0.36-0.74). This was in contrast to the high discriminative performance of the smoking scores (internal AUC 0.80, 0.69-0.91 and external AUC 0.80, 0.72-0.88; **Supplementary Figure 4**). The DNA methylation scores for both smoking and e-cigarette use performed poorly at discriminating tumour from adjacent normal tissue in lung adenocarcinoma (LUAD) cases (AUC 0.57, 0.40-0.73 and 0.58, 0.42-0.74). The e-cigarette score also performed poorly at discriminating tumour from adjacent normal tissue in lung squamous cell carcinoma (LUSC) cases (0.64, 0.52-0.76). Moderate discrimination was observed from the smoking score in LUSC cases (0.73, 0.61-0.85) (**Supplementary Figure 5**).

## Discussion

Using saliva samples from 117 smokers, 117 non-smokers and 116 vapers with a limited smoking history, we found that the DNA methylation signals of e-cigarette use were weak and largely distinct from those established in relation to cigarette smoking. In particular, a lack of enrichment for previous smoking-related CpG sites was observed in the vapers compared with the smokers. We found modest evidence for differential methylation related to e-cigarette use at a number of sites or regions proximal to genes related to metabolism (*IGF1R, SLC2A10, THRB*), cancer (*IGF1R, BLCAP, LINC00857*) and the nervous system (*NNAT, ANXA11, GABRP*).

Vapers also exhibited differential methylation relative to non-smokers in *MUC4* encoding Mucin 4, an integral membrane glycoprotein present in mucus which has previously been found to be upregulated in vapers.^39^ Ethanol/alcohol, positive regulation of insulin secretion, GABA transport and butanoate metabolism were among the most enriched pathways, which may reflect biological responses to e-cigarette constituents (ethyl alcohol, ethyl butyrate and nicotine).

Two previous studies (comprising 32 and 45 participants, respectively) have found associations between e-cigarette use and DNA methylation levels which overlapped with smoking-related signals.^27 40^ This is in contrast to the present study where DNA methylation scores for smoking were similar between vapers and non-smokers. We were also unable to replicate DNA methylation differences for 14 CpGs previously related to e-cigarette use^27^. Previous findings may therefore represent false positives given previously small sample sizes or residual confounding by smoking status. Alternatively, the lack of replication could be due to tissue differences, since the previous study assessed methylation in bronchoalveolar samples.

We also investigated DNA methylation scores for biological ageing, and found that, while these were again similar between vapers and non-smokers, higher levels of a biological ageing score (GrimAge) were observed in smokers. This is of interest since epigenetic age is predictive of age-related disease and mortality independent of chronological age,^41-43^ and so may serve as a predictor of higher mortality rates among smokers.

Finally, a DNA methylation score generated to index e-cigarette use poorly discriminated vapers from non-smokers in SEE-Cigs and in an independent dataset (ALSPAC). This was in contrast to a DNA methylation score generated to index smoking, which discriminated smokers from non-smokers in both datasets. The smoking score also showed better discrimination of tumour and adjacent normal tissue in lung squamous cell cases compared with the e-cigarette score. These findings are supported by a previous study which found that a DNA methylation profile for smoking derived from oral samples was able to discriminate tumour and adjacent normal samples with high accuracy.^14^ This indicates that DNA methylation scores for smoking may incorporate more epigenetic aberrations relevant to tumourigenesis than an equivalent score for e-cigarette use. However, the smoking DNA methylation score generated in the present study was lower in lung squamous cell carcinoma relative to normal tissue, the inverse of what was expected due to higher levels being observed in smokers.^14^ While this is surprising, we have previously found differential methylation in tumour tissue is in the opposite direction to that observed in relation to smoking for *AHRR* (cg05575921)^*44*^ (**Supplementary Figure 5**), the CpG contributing most weight to the DNA methylation score for smoking (**Supplementary Table 18**).

Major strengths relate to the design of the study, including the recruitment individuals with a limited smoking history and the assessment of DNA methylation levels in an easily accessible and exposure-relevant tissue in order to investigate epigenetic profiles of e-cigarette use. Limitations include the representativeness of our study sample, with demographic characteristics different to the general population due to the strict inclusion criteria. The young age of the study sample (mean age = 21 years) and limited smoking and vaping history could hamper the detection of DNA methylation signals. This may explain why so few CpG sites were identified at epigenome-wide significance in relation to smoking than anticipated based on previous studies of oral samples^14^. Nonetheless, we found evidence for enrichment of smoking-related CpG sites among the smokers, suggesting that the signals were present but below the strict Bonferroni-correction threshold. Similar enrichment was not found among the vapers, indicating that the methylation signature of e-cigarette use is distinct from that of smoking, and that vapers in the present study were accurate in their reporting of having a limited smoking history, despite this not being biochemically verified.

Additional research in cohort studies is required to investigate DNA methylation changes among ex-smokers quitting with different methods, including e-cigarettes. It would be of particular interest to investigate dual users in comparison with exclusive smokers and vapers to assess whether methylation signals are more prominent in this group, who make up the majority of e-cigarette users. While findings from this study suggest that e-cigarettes may have distinct health effects from cigarettes, we cannot provide robust conclusions regarding the safety of e-cigarettes. Furthermore, although the DNA methylation changes identified in relation to both smoking and e-cigarette use may be predictive of future disease risk, the causal consequences of these DNA methylation changes on health outcomes are currently uncertain.^44^

Our study was powered to detect an 11% mean difference in DNA methylation at p<10^−6^. This is below that typically required for obtaining epigenome-wide significant findings, although is adequate for ensuring replication of previously-associated CpGs, such as that at *AHRR* (cg05575921) in relation to smoke exposure, which has been found to exhibit up to 40% higher DNA methylation in current versus never smokers in both blood and saliva samples^17^. Therefore, while we can be confident in our finding that differential methylation at this site is specific to smokers and not vapers, we were less powered to detect novel CpGs in relation to e-cigarette use. While it appears that the DNA methylation profile of vapers is less pronounced than that of smokers, the DNA methylation changes associated with e-cigarette use may be commensurate in scale with other lifestyle exposures and replication of the signals identified in relation to e-cigarette use in larger studies is warranted.

## Conclusions

Findings from this study suggest that e-cigarette use does not impact saliva DNA methylation in the same way as cigarette smoking. Unlike for smoking, the DNA methylation profile for e-cigarette use did not replicate in independent samples and was not able to discriminate cancer from normal tissue. Further studies are required to detect a robust methylation signature for long-term e-cigarette use. The extent to which differential methylation related to e-cigarette use translates into chronic effects and relevant health outcomes should also be investigated.

## Data Availability

Results from the epigenome-wide association studies performed will be uploaded to the EWAS catalog (http://www.ewascatalog.org/)

## Acknowledgements

**SEE-Cigs** – We are extremely grateful to all the participants who took part in this study.

**ALSPAC** - We are extremely grateful to all the families who took part in this study, the midwives for their help in recruiting them, and the whole ALSPAC team, which includes interviewers, computer and laboratory technicians, clerical workers, research scientists, volunteers, managers, receptionists and nurses.

## Author contributions

Study conception and design: R.C.R., G.S., M.S., C.L.R., M.M., S.H.G.

Data acquisition, analysis and interpretation of data: R.C.R., C.S.R., J.N.K., C.P., A.B., S.H.G.

Drafting the manuscript: R.C.R., C.S.R., S.H.G.

Critically revising the manuscript: J.N.K., C.P., A.B., G.S., M.S., C.L.R., M.M.

Final approval: All authors

## Funding

This work was supported by a Cancer Research UK Population Research Committee project grant (C57854/A22171). Further support was provided by the UK Medical Research Council (MC_UU_00011/5 and MC_UU_00011/7), which funds a Unit at the University of Bristol where R.C.R., C.S.R., J.K., C.P., G.S., M.S., C.L.R and M.M. work, and the CRUK-funded Integrative Cancer Epidemiology Programme (C18281/A19169). R.C.R is a de Pass Vice Chancellor’s Research Fellow at the University of Bristol. C.S.R’s time is supported by the National Institute for Health Research Applied Research Collaboration West (NIHR ARC West) at University Hospitals Bristol NHS Foundation Trust. The UK Medical Research Council and Wellcome (Grant ref: 102215/2/13/2) and the University of Bristol provide core support for ALSPAC. This publication is the work of the authors who will serve as guarantors for the contents of this paper. A comprehensive list of grants funding is available on the ALSPAC website (http://www.bristol.ac.uk/alspac/external/documents/grant-acknowledgements.pdf).

## Competing interests

None to declare.

## Notes

### Competing Interest Statement

The authors have declared no competing interest.

### Author Declarations

Ethics approval for the study was granted by the Faculty of Science Human Research Ethics Committee at the University of Bristol

