## Supplementary material for "Investigating the DNA methylation profile of e-cigarette use": Online Supplement

Supplementary Methods

Eligibility criteria

In order to maximise the chance that vapers had never been cigarette smokers, and to minimize confounding by age, we restricted eligibility to between 16 and 35 years old. Additional inclusion criteria were that the participants were in good physical and mental health (measured via self-report) and were able to give informed consent as judged by the investigator. Exclusion criteria obtained via self-report were: dependence on alcohol or drugs (other than nicotine); significant current or past illness (including cancer and type 1/type 2 diabetes); current pregnancy or breast feeding; having a related individual in the sample (1).

Recruitment

Participants were recruited via a number of mechanisms including from the student population at the University of Bristol, podcasts, blogs, posters/flyers in vape shops, and social media. Recruitment began in January 2017 and was completed in January 2019. The study protocol was originally published on the Open Science Framework on 19/01/2017. On 06/02/2018 we were granted ethics approval to relax the eligibility criteria related to age (16-35 years) and previous smoking history of the vapers and never-smokers (<100 cigarettes in their lifetime) and vaping history of the smokers and never-smokers (vaped <100 times in their lifetime), which were initially <50. We stated in our original protocol that we would relax age criteria if recruitment stalled. Ethics approval for the study was granted by the Faculty of Science Human Research Ethics Committee at the University of Bristol. All participants have provided written informed consent.

Questionnaire

Based on initial responses to the questionnaire, and in accordance with the eligibility criteria, participants were allocated to three participant groups (smokers, vapers and never-smokers). Participants in the ‘smokers’ category were asked whether they smoke cigarettes or roll-ups, whether they were daily or weekly smokers, and how many cigarettes they smoked per day/week as appropriate. They were asked at what time of day they smoke their first cigarette, for how long they had been a smoker, and details about whether they had plans to give up, or whether they had previously attempted to stop smoking. Participants in the ‘vapers’ category were asked the type of device they used, the nicotine concentration they used most frequently, and whether they had changed the nicotine concentration in the past. If they reported using a refillable device, they were asked to estimate the volume of liquid used in an average day. Similarly to the smokers group this group were asked the time of day that they first vape, how long they have been a vaper, and details about whether they had plans to give up vaping. Participants in the ‘never-smokers’ group were asked whether they had ever smoked or vaped, and how frequently, to ensure they met our inclusion/exclusion criteria.

Sample collection

After completing an online questionnaire, participants were screened for eligibility and sent an information sheet and consent form. On enrolling, participants were posted a study pack containing a saliva collection (DNA Genotek Oragene™) kit from which DNA was extracted and methylation was measured. We supplemented existing kit instructions with a simplified version to aid understanding and improve sample quality, which was posted to participants along with the kit, consent form and information sheet. We asked participants to provide 2 mL of saliva and return the kits through the post to the University of Bristol, where they were processed by the Bristol Bioresource Laboratories.

Sample processing, data quality control and normalization

DNA was extracted from the saliva samples and underwent bisulphite conversion using the Zymo EZ DNA Methylation^TM^ kit (Zymo, Irvine, CA). DNA samples were loaded onto the Illumina HumanMethylationEPIC array in three batches with sampling criteria in place to ensure that all three groups were represented in each batch in order to minimise potential confounding by batch effects. In addition, during the data generation process a wide range of batch variables were recorded in a purpose-built laboratory information management system (LIMS), which also reported quality control (QC) metrics. Microarray data underwent quality control and normalization using *meffil*, an R package designed for pre-processing of large samples of Illumina Methylation BeadChip microarrays (2). Sample outliers were identified and removed based on sex-chromosome methylation, methylation vs unmethylation intensity, control probes, detection p-values (N=10 exclusions in total: 4 vapers, 3 smokers and 3 non-smokers). Poor quality CpG sites, SNP/control probes and CpGs on the sex chromosomes were excluded, resulting in 846,244 CpG sites for analysis.

Estimated cell type proportions

A cell type reference for saliva was derived as part of *meffil* by combining a white blood cell type reference (GEO: GSE35069) and a buccal cell type reference (GEO: GSE48472). Estimated cell type proportions comprised: Buccal, CD4T, CD8T, Monocytes, B-cells, NK cells and Granulocytes.

DNAm scores of smoking and epigenetic ageing

We assessed associations between DNAm scores comprising methylation values derived from a weighted average of CpG sites found to be related to smoking in previous studies. This included scores derived from the CpG sites identified in EWAS conducted by Joehanes et al. (3) and Teschendorff et al. (4), as well as 233 and 172 CpG sites identified in McCartney et al. (5) and Lu et al. (6), respectively. The latter two studies used penalised regression models of smoking pack-years to identify CpG sites most predictive of smoke exposure. Finally, since the CpG site, cg05575921 (*AHRR*) contributed most weight to all of the DNAm scores and has been proposed as an independent biomarker of smoking (7), we investigated this sites as an additional biomarker. With the exception of *AHRR*, the other scores developed were linear combinations of methylation levels at the relevant CpG sites weighted by the effect sizes of sites identified in relation to smoking from the various studies (3-6).

For epigenetic ageing, we assessed associations between two “first generation” epigenetic clocks derived from DNAm levels at CpG sites found to be strongly associated with chronological age (8, 9), as well as two more recently derived clocks: one optimised to predict physiological dysregulation (PhenoAge) (10) and one optimised to predict lifespan (GrimAge) (6). To generate the epigenetic ageing measures in SEE-Cigs, we uploaded DNAm data for a subset of CpG sites from the Illumina EPIC array to the online DNAm Age Calculator https://dnamage.genetics.ucla.edu/ developed by the Horvath lab. We also uploaded an annotation file, containing data on chronological age, sex and tissue type (saliva) for the samples. We were able to generate the following epigenetic ageing measures: intrinsic epigenetic age acceleration based on Horvath’s multi-tissue predictor (IEAA) (8); extrinsic epigenetic age acceleration (EEAA) based on Hannum’s method, which up-weights the contribution of blood cell composition (11); PhenoAge (10), GrimAge (6). Intrinsic epigenetic age acceleration (IEAA) is independent of changes in blood cell composition while extrinsic epigenetic age acceleration (EEAA) incorporates age-related changes in blood cell composition. PhenoAge and GrimAge can be considered as measures of extrinsic ageing.

DNAm scores of e-cigarette use

*SEE-Cigs*

For internal validation of the DNAm score of e-cigarette use, we used a training (2/3 sample of vapers and non-smokers) and testing set (1/3 sample of vapers and non-smokers) within the SEE-Cigs study. In the training set, we used the *glmnet* package in R to fit a generalized logistic regression via penalized maximum likelihood using three-fold cross validation and run 10 times to determine a lambda with minimum average error. A DNAm score was then generated based on the fitted object produced. The resulting DNAm score comprised a sum of the beta-values of the included CpG sites. Its performance in predicting e-cigarette use was evaluated in the test stet by generating a receiver operator characteristic (ROC) curve and evaluating the area under the curve (AUC) derived from the logistic regression model using the R package *pROC* (version 1.16.1)*.* We compared the AUC obtained from this model with that from a similar model for predicting smoking. For this, we derived a DNAm score for smoking using a training set (2/3 sample of smokers and non-smokers) and testing set (1/3 sample of smokers and non-smokers) within SEE-Cigs.

*Avon Longitudinal Study of Parents and Children*

We next assessed external validation of DNAm scores for e-cigarette use and smoking in the Avon Longitudinal Study of Parents and Children (ALSPAC). ALSPAC is a large, prospective cohort study based in the south-west of England. Pregnant women resident in Avon, UK with expected dates of delivery 1^st^ April 1991 to 31^st^ December 1992 were recruited and detailed information has been collected on these women and their offspring at regular intervals (12, 13). Additional offspring that were eligible to enroll in the study have been subsequently recruited at the ages of 7 and 18 years (14). The additional enrolment provides a baseline sample of 14,901 offspring who were alive at 1 year of age. Please note that the study website contains details of all the data that are available through a fully searchable data dictionary and variable search tool (<http://www.bristol.ac.uk/alspac/researchers/our-data>). Ethical approval for the study was obtained from the ALSPAC Ethics and Law Committee and the Local Research Ethics Committees. Consent for biological samples has been collected in accordance with the Human Tissue Act (2004). Informed consent for the use of data collected via questionnaires and clinics was obtained from participants following the recommendations of the ALSPAC Ethics and Law Committee at the time,

When the offspring were 24 years old, they were invited to attend the Focus @ 24+ clinic, which took place between June 2015 and October 2017. 4,026 were seen at this clinic, where fasting blood samples were taken. Blood samples from 570 of the offspring were selected for DNA methylation profiling to maximize overlap with existing DNA methylation profiles generated collected at younger ages as part of the Accessible Resource for Integrated Epigenomic Studies (ARIES) (15). Following DNA extraction, samples were bisulfite-converted using the Zymo EZ DNA MethylationTM kit (Zymo, Irvine, CA). Genome-wide methylation was then measured using Illumina Infinium MethylationEPIC Beadchip arrays. The arrays were scanned using an Illumina iScan, with initial quality review using GenomeStudio. During the data generation process a wide range of batch variables were recorded in a purpose-built laboratory information management system (LIMS), which also reported quality control metrics. Quality control and normalization was then carried using the *meffil* R package (2). Quality control included checks for sample swaps using genotype matching and sex prediction, methylated vs unmethylated signal outliers, dye bias, poor probe signal detection, and low bead numbers. Only one sample failed and was excluded due to evidence of being a sample swap and having multiple control probe outliers. The 569 samples that passed were normalized in *meffil* using functional normalization using the top 20 control probe principal components and sample plate as a random effect.

At the same time point when the offspring were 24 years old, information on smoking and e-cigarette use was obtained from a questionnaire that was completed by 458 of the offspring with DNA methylation data. Questionnaire data were collected and managed using REDCap electronic data capture tools hosted at the University of Bristol (16). The participants were asked whether they had ever smoked or used an e-cigarette, as well as about the frequency and duration of use. From these details were determined three groups of participants: 1) vapers (currently users of electronic cigarettes or other vaping devices, n=14) 2) smokers (current daily or weekly smokers, n=47) 3) non-smokers (smoked <100 cigarettes in their lifetime and never used an electronic cigarette or other vaping device, n=262).

We assessed the discriminative performance of a DNAm score generated in SEE-Cigs for predicting e-cigarette use (vs non-smoking) in the ALSPAC cohort, which was compared with a DNAm score for predicting smoking (vs non-smoking). Both e-cigarette and smoking DNAm scores were obtained using the same approach described above in the full sample of: i) vapers vs non-smokers and ii) smokers vs non-smokers, respectively, in SEE-Cigs.

*The Cancer Genome Atlas*

We used data on 27 individuals with lung adenocarcinoma (LUAD) and 41 individuals with lung squamous cell carcinoma (LUSC) who had Illumina Infinium 450K DNAm measured in both tumour and adjacent normal samples as part of The Cancer Genome Atlas (TCGA). Again, a DNAm score was generated on the full sample of vapers vs. no-smokers in SEE-Cigs, this time restricted to CpG sites which were present only on the 450K array. This was compared with a DNAm score for smoking generated on the full sample of smokers vs. non-smokers, again restricted to 450K CpG sites.
