## Supplementary Table for "Investigating the DNA methylation profile of e-cigarette use"

**Supplementary Figure 1** – Analysis plan

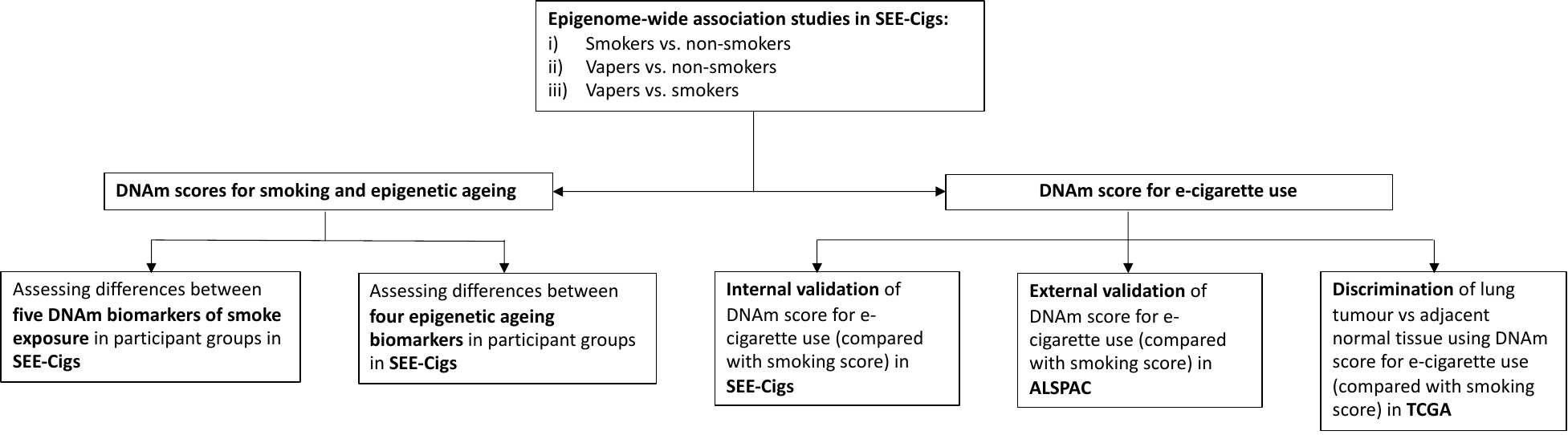

**Supplementary Figure 2:** Heatmap of differentially methylated CpG sites identified among smokers, vapers and non-smokers

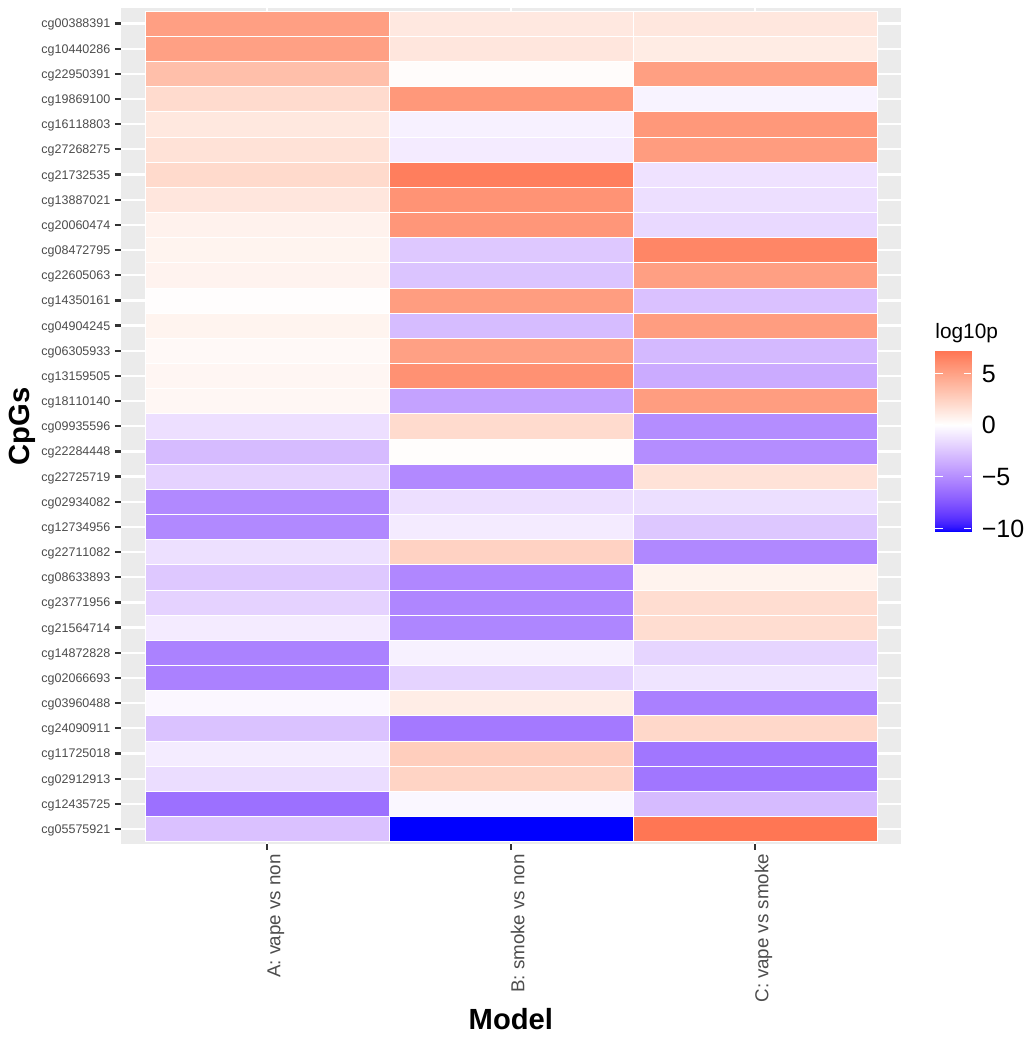

**Supplementary Figure 3** – Enrichment of known smoking related CpG sites in SEE-Cigs

A)	B)

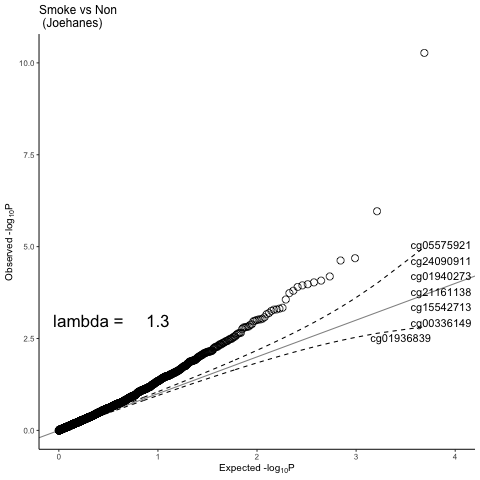

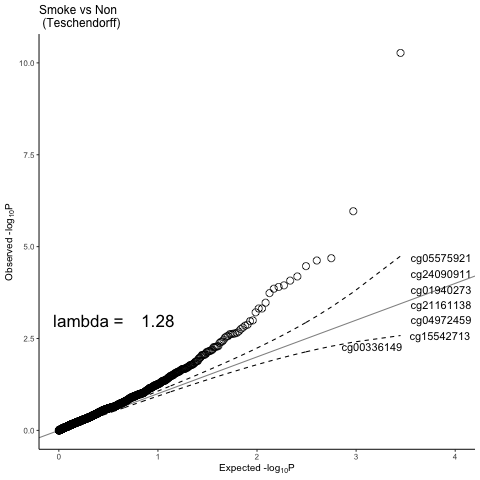

C)	D)

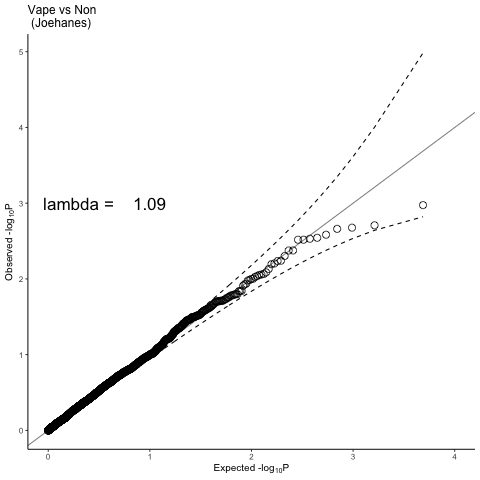

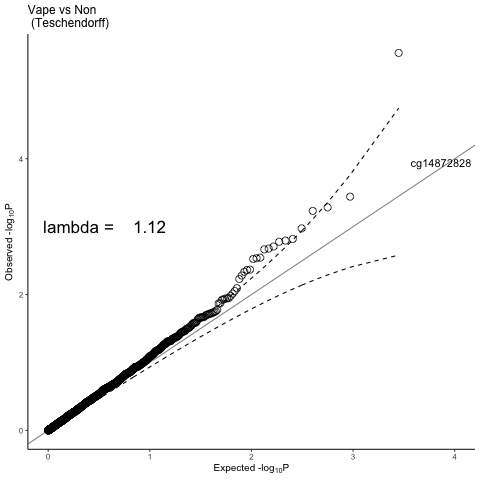

A) CpG sites from Joehanes et al (*1*) in EWAS of smokers vs non-smokers in SEE-Cigs
B) CpG sites from Teschendorff et al (*2*) in EWAS of smokers vs non-smokers in SEE-Cigs
C) CpG sites from Joehanes et al (*1*) in EWAS of vapers vs non-smokers in SEE-Cigs
D) CpG sites from Teschendorff et al (*2*)in EWAS of vapers vs non-smokers in SEE-Cigs

**Supplementary Figure 4 -** Comparing the discriminative performance of a DNAm score for e-cigarette use with a DNAm score for smoking in independent samples

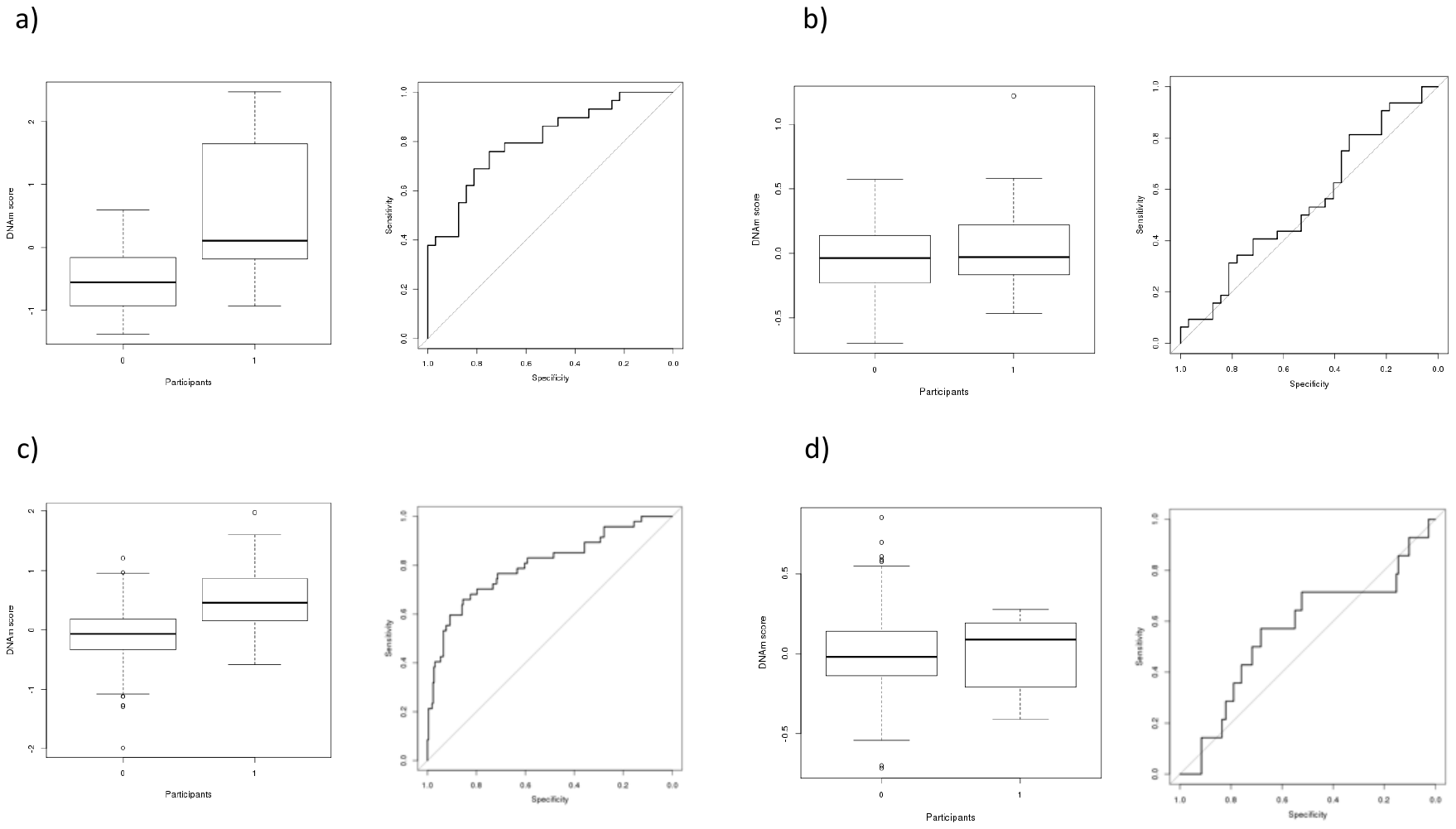

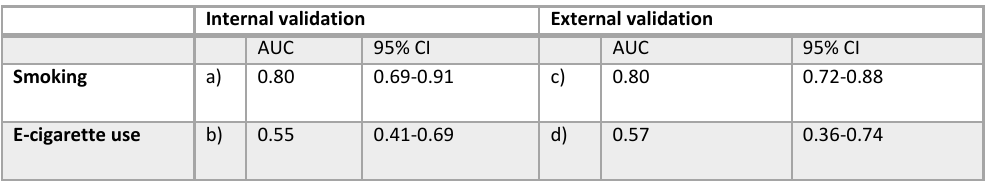

a) Performance of a DNAm score for smoking in discriminating smokers (1) from non-smokers (0) in SEE-Cigs (n= 32 smokers and n= 32 non-smokers)
b) Performance of a DNAm score for e-cigarette use in discriminating vapers (1) from non-smokers (0) in SEE-Cigs (n= 32 vapers and n= 32 non-smokers)
c) Performance of a DNAm score for smoking in discriminating smokers (1) from non-smokers (0) in ALSPAC (n= 47 smokers and n= 262 non-smokers)
d) Performance of a DNAm score for e-cigarette use in discriminating vapers (1) from non-smokers (0) in ALSPAC (n= 14 vapers and n= 262 non-smokers)

**Supplementary Figure 5 -** Comparing the performance of a DNAm score for e-cigarette use with a DNAm score for smoking in discriminating lung tumour from normal adjacent tissue

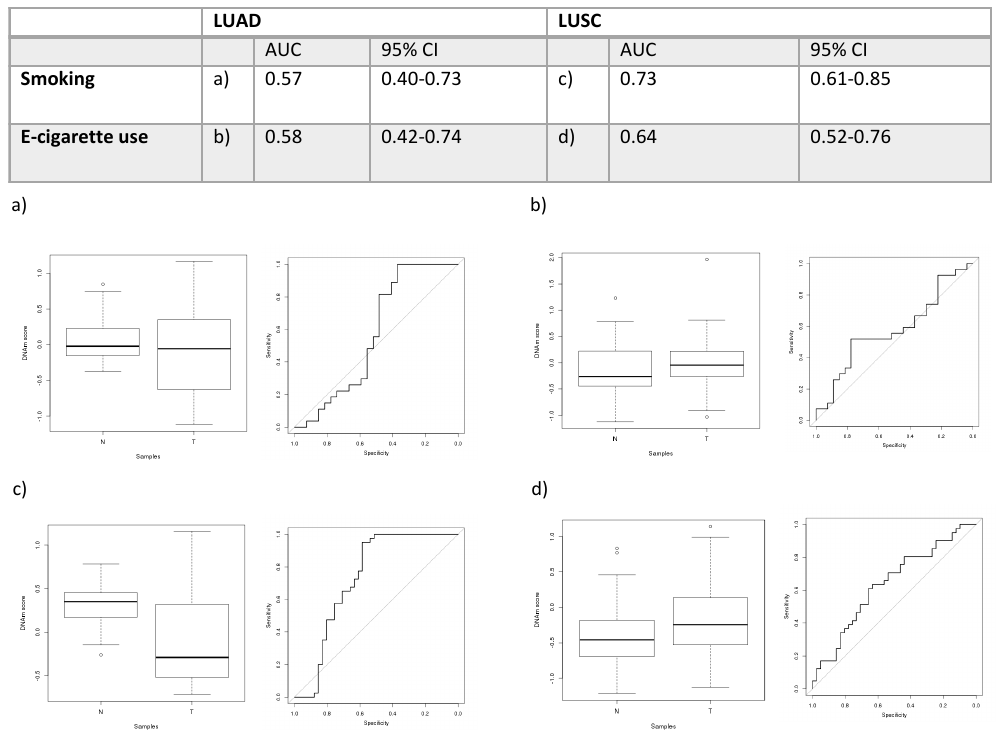

a) Performance of a DNAm score for smoking in discriminating lung adenocarcinoma (LUAD) tumour (T) from adjacent normal (N) in TCGA (n= 27 matched pairs)
b) Performance of a DNAm score for e-cigarette use in discriminating lung adenocarcinoma (LUAD) tumour (T) from adjacent normal (N) in TCGA (n= 27 matched pairs)
c) Performance of a DNAm score for smoking in discriminating lung squamous cell carcinoma (LUSC) tumour (T) from adjacent normal (N) in TCGA (n= 40 matched pairs)
d) Performance of a DNAm score for e-cigarette use in discriminating lung squamous cell carcinoma (LUSC) tumour (T) from adjacent normal (N) in TCGA (n= 40 matched pairs)

**Supplementary Figure 6** – Assessing the discriminative performance of AHRR (cg05575921) methylation

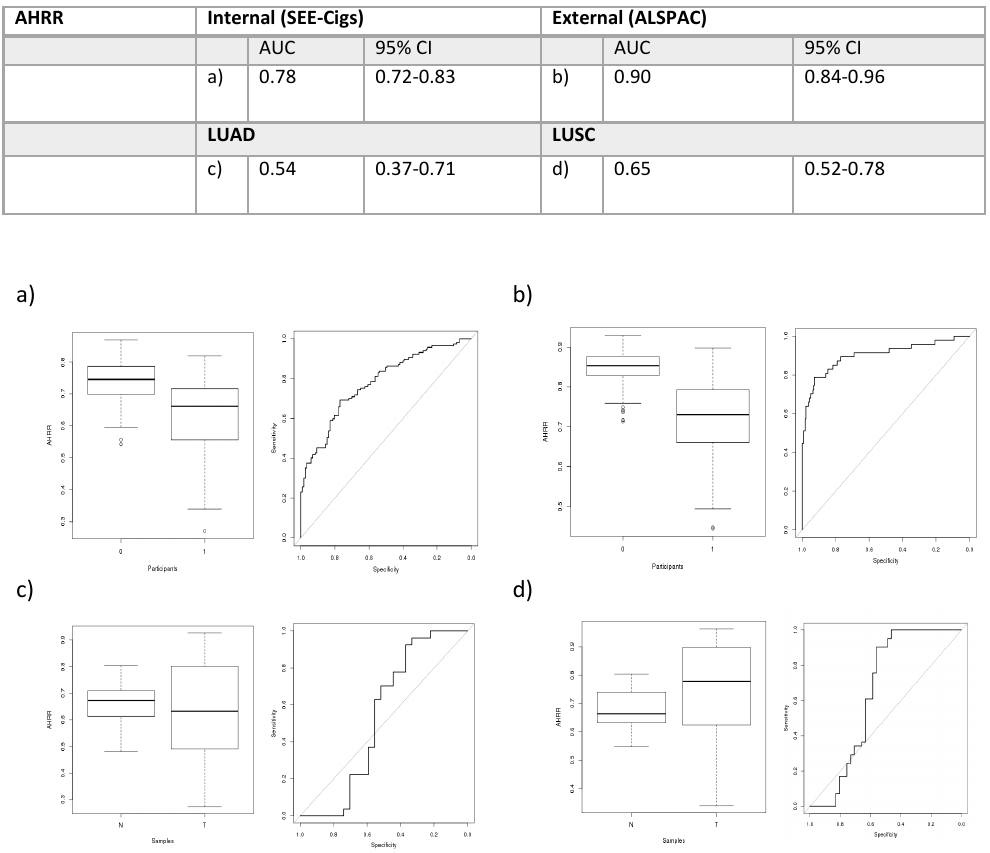

a) Performance of *AHRR* (cg05575921) methylation in discriminating smokers (1) from non-smokers (0) in SEE-Cigs (n= 32 smokers and n= 32 non-smokers)
b) Performance of *AHRR* (cg05575921) methylation in discriminating smokers (1) from non-smokers (0) in ALSPAC (n= 47 smokers and n= 262 non-smokers)
c) Performance of *AHRR* (cg05575921) methylation in discriminating lung adenocarcinoma (LUAD) tumour (T) from adjacent normal (N) in TCGA (n= 27 matched pairs)
d) Performance of *AHRR* (cg05575921) methylation in discriminating lung squamous cell carcinoma (LUSC) tumour (T) from adjacent normal (N) in TCGA (n= 40 matched pairs)

1.	R. Joehanes *et al.*, Epigenetic Signatures of Cigarette Smoking. *Circ Cardiovasc Genet* **9**, 436-447 (2016).

2.	A. E. Teschendorff *et al.*, Correlation of Smoking-Associated DNA Methylation Changes in Buccal Cells With DNA Methylation Changes in Epithelial Cancer. *JAMA Oncol* **1**, 476-485 (2015).
